# Caffeine decreases neuromuscular fatigue in the lumbar muscles – a randomized blind study

**DOI:** 10.1101/2020.06.08.20122531

**Authors:** Liziane Cardoso, Tatyana Nery, Maielen Gonçalves, Maria Carolina Speck, Ana Cristina de Bem Alves, Viviane Bristot, Thais Gonçalves, Débora da Luz Scheffer, Ione Jayce Ceola Schneider, Heloyse Kuriki, Aderbal S Aguiar

## Abstract

**Background:** Ergogenic evidence for caffeine is robust in sports and isolated limb tasks. Our objective was to evaluate a possible ergogenic effect on postural muscles, still unknown, through the Biering-Sørensen’s lumbar extension test.

**Methods:** A double-blind, controlled placebo, crossover study. Fifty-one healthy, physically inactive male subjects (18-25 years) with average body mass (BMI 18.5 – 24.9 kg/m^2^) were recruited. The subjects received oral caffeine (6 mg/kg) and saline (0.3%) in two cross-evaluations separated by one week. The primary outcome was the time in the Biering-Sørensen test after 1 hour of treatment. The secondary outcomes were peak lumbar extension force, rating of perceived exertion, EMG’s median frequency and muscle recruitment of *multifidus spinae and transversalis*/interne oblique muscles, and cardiovascular variables (heart rate and blood pressure).

**Results:** 27 subjects were blindly treated with caffeine and saline. Caffeine was ergogenic during the Biering-Sørensen test. It increased lumbar extension time (*d* = 0.34, P<0.05), but not peak force. The perception of effort decreased with caffeine (*d* = 0.37, P<0.05). Caffeine increased muscle stimulation frequency (P<0.05) and recruitment (*η^2^* = 0.49, P<0.05) of *multifidus spinae*. In the transversalis/interne oblique muscles, caffeine increased the median frequency *η^2^* = 0.13, P<0.05) and the distribution of higher frequencies (P<0.05). Caffeine also increased muscle recruitment in the transversalis/interne oblique muscles *η^2^* = 0.13, P<0.05) Tachycardia and increased blood pressure at the lumbar test were higher in the caffeine condition (P< 0.05).

**Conclusions:** Our results show that caffeine is ergogenic for postural muscles. Decreased RPE and improved muscle activity suggest central mechanisms of caffeine.

**Trial registration:** UTN U1111-1234-2079.

**BULLET POINTS:**

1. Caffeine increases the extension time of the lumbar spine in the Biering-Sørensen test.
2. Caffeine decreases perceived exertion during the Biering-Sørensen test.
3. Caffeine increases muscle stimulation and recruitment of the multifidus spinae during the Biering-Sørensen test.
4. Caffeine increases muscle stimulation of the transversalis/interne oblique muscles during the Biering-Sørensen test.

## INTRODUCTION

Caffeine (1,3,7-trimethylxantine) is one of the most used ergogenic substances by physical activity practitioners and athletes [1–10]. Caffeine is hydrophilic, fast absorbed, and distributed to all body tissues [11,12]. Caffeine, a non-selective competitive adenosine receptor antagonist (A_1R_ and A_2A_R subtypes), increases endurance [1,8–10,13,14], intermittent [7,15,16] and resistance [4,17] exercise. The most used caffeine dose is 6 mg/kg, 1-hour before physical activity [1–10]. Ergogenic doses of caffeine have minor adverse effects, such as angiogenesis, insomnia, and gastrointestinal discomfort [18].

The main pharmacological effect is to counteract adenosine, an inhibitory modulator of the central nervous system (CNS) [11,12]. Evidence suggests that caffeine modifies neurological activity when ergogenic. Caffeine decreases rate of perceived exertion [4–6,9,19], pain [4,6,20–22], central and mental fatigue during exercise[23–26]. Studies are showing decreased exercise-induced saccade fatigue [23,24], increased brain motor area potentiation after transcranial magnetic stimulation [25], and modifies spinal excitability and cortical silence in fatigued muscles [27,28]. Caffeine also improves exercise performance expectations [26], cognitive and executive functions [19,29–31], and vigor [32] in exercising subjects. In animal studies, caffeine fails to be ergogenic in a mouse lacking neuronal A_2A_R [33] and rats treated with 5’-N-ethylcarboxamidoadenosine (NECA), an adenosine analog [34,35].

We conducted this study to evaluate the effects of caffeine on endurance (Biering-Sørensen test) and electrical activity (electromyography – EMG) of core muscles, a well-controlled experiment in our laboratory [36]. There is ergogenic evidence for upper and lower limb muscles [37–40], but not for core muscles. Recently, these muscles are part of the sports training routine, as they are essential for performance enhancement [41,42] and injury prevention [43,44] in athletes. The possible ergogenic effects of caffeine on core muscles can contribute to better periodization, as well as used in regeneration and treatment mesocycles, in addition to dedicated mesocycles for performance, tapering, and competitions.

## METHODS

### Study design and subjects

We conducted a randomized, double-blind, placebo-control, crossover study, where both examiner and subjects were unaware of ingested substances. Recruitment was carried out through social networks in the community within the micro-region of Araranguá (south latitude 28º56’05 “and west longitude 49º29’09”), southern Brazil. Subjects also collaborated in the recruitment of others, according to the public involvement statement. We called the volunteers for clarification on the study design, and the inclusion and exclusion criteria. The study followed the CONsolidated Standards of Reporting Trials (CONSORT).

We recruited healthy, physically inactive male subjects with average body mass (BMI 18.5 – 24.9 kg/m^2^) aged 18–25 years. The screening included a semi-structured interview about medical regarding medical history, coffee consumption [45], and the International Physical Activity Questionnaire (IPAQ) – short form – Brazilian version [46–48]. Exclusion criteria: smokers; hypertension; diabetes; lumbar and orthopedic diseases; neurological, cardiac and gastric diseases; continued use of antidepressants and medications or supplements containing caffeine; and engaging physical activities more than two times/wk.

The study respected the declaration of Helsinki. Moreover, the Brazilian National Health Council also requires compliance with ethical principles of Resolution No. 466 of December 12, 2012. All subjects signed a consent form before inclusion in the study. The trial was approved by the Brazilian Ethics Committee (http://plataformabrasil.saude.gov.br) with file number CAEE 86400418.0.0000.0121 and was prospectively registered (trial RBR-39JNP4) in the Brazilian Clinical Trials Register (http://www.ensaiosclinicos.gov.br/). The Universal Trial Number (UTN) is U1111-1234-2079.

### Randomization and blinding

All eligible subjects were evaluated when treated with vehicle or caffeine, blinded to the researchers through tubes reporting substance 1 or A, respectively. Subjects were randomized in a 1:1. One independent researcher (MCS) carried out simple randomization with a coin toss to start the test with a blinded substance. Other blinded research assistants (LC, TN, and MG) to treatment administered all outcome measures. The statistician was unaware of treatment allocation before completion of data collection and data cleaning.

### Intervention

Caffeine and sodium chloride (NaCl) with 98% purity was purchased from Sigma-Aldrich® (Brazil). The subjects’ body mass was measured, a concentrated caffeine solution (3 mg/mL) was prepared freshly, and the individual volume adjusted by multiplying body mass by two. The vehicle was a 0.3% NaCl in tap water. Subjects drank identical volumes of caffeine (6 mg/kg) or vehicle, 1-hour before the Biering-Sørensen test, the time necessary to reach plasma peaks. A researcher (LC) followed the subjects for 24 hours to monitor the development of adverse effects of caffeine or other symptoms.

### Measurements

The analyzes were performed in the Evaluation and Rehabilitation of the Locomotor System Lab (Laboratório de Avaliação e Reabilitação do Aparelho Locomotor – LARAL, UFSC, Araranguá-SC, Brazil). Participants were advised to continue habitual eating throughout the study. Physical activity, alcohol consumption, and caffeine (food and beverages) were avoided in the 48 hours before the tests. Each test was performed on two separate days, with 1-week intervals, during the morning or afternoon, according to the subjects’ agenda availability. The researchers sought all subjects for evaluations in the laboratory.

After 1 hour of oral treatment, the endurance of *multifidus spinae* was assessed by peak extensor force [36], and the Biering-Sørensen test [36]. At the same time, electromyography (EMG) assessed the median frequency and muscle recruitment (Root mean Square of EMG signal – RMS) of multifidus spinae and transversalis/interne oblique muscles [36], Borg Rating of Perceived Exertion – RPE – Brazilian version (30-s intervals) [49,50] were performed. Heart Rate (HR) was assessed during rest and at 15-s intervals during the test. At the end of the test, we evaluated lactate capillary blood (earlobe) levels [51,52] and collected antecubital venous blood samples for plasma separation and storage (−80°C). Plasma caffeine concentrations were assessed using High-Performance Liquid Chromatography – HPLC [53].

### Outcomes and sample size

We designed this trial to detect changes in the Biering-Sørensen test time (primary outcome) and peak force (secondary outcome), RPE (secondary outcome), and EMG’s median frequency and muscle recruitment (secondary outcome). The study was carried out with 27 subjects to have 85% power with alpha = 0.05.

### Statistics

A blinded researcher (ASAJr) performed the statistical analysis according to an intention-to-treat principle. The results were described on mean ± SD. We assessed the normal distribution of results before statistical comparisons. Paired Student’s t-test analyzed Biering-Sørensen time and peak force, blood lactate levels, RPE and HR (area under the curve – AUC), and final 10% of EMG median frequency. The final 10% of the RMS was assessed through Wilcoxon matched-pairs signed-rank test. This analysis was performed using GraphPad Prism (v. 5.0, GraphPad Software). The STATISTICA (v. 8.0, StatSoft, Inc.) was used to compare ANOVA of EMG median frequency and RMS, chi-square test of EMG median frequencies, and Mann Whitney test of HR. P< 0.05 was considered significant. Effect sizes (Cohen’s d) were calculated for between-group changes in mean differences for all outcome measures, where a Cohen’s *d* = 0.2 represents a ‘small’ effect size, 0.5 represents a ‘medium’ effect size and 0.8 a ‘large’ effect size [54]. Cohen’s *η*^2^ was used for repeated measurements, defined as 0.02 small, 0.13 medium, and 0.26 large [54].

### Public Involvement statement

There were no funds or time allocated for public involvement, so we were unable to involve subjects. We have invited subjects to help us develop our dissemination strategy.

## RESULTS

### Subjects and plasma caffeine levels

Fifty-one subjects were recruited between 7/15/2018 and 12/15/2018, interrupted on 5/11/2018 when the sample size was completed. Fig. 1 shows the participant flow. Figure 1 shows the trial profile as per the Consolidated Standards of Reporting Trials [55]. There were 27 undergraduate students, single, with ideal body mass index, irregularly active and light (25.9%), moderate (48.1%), and high (25.9%) coffee consumers. Table 1 shows the subjects’ demographic and baseline data.

**Fig. 1.**
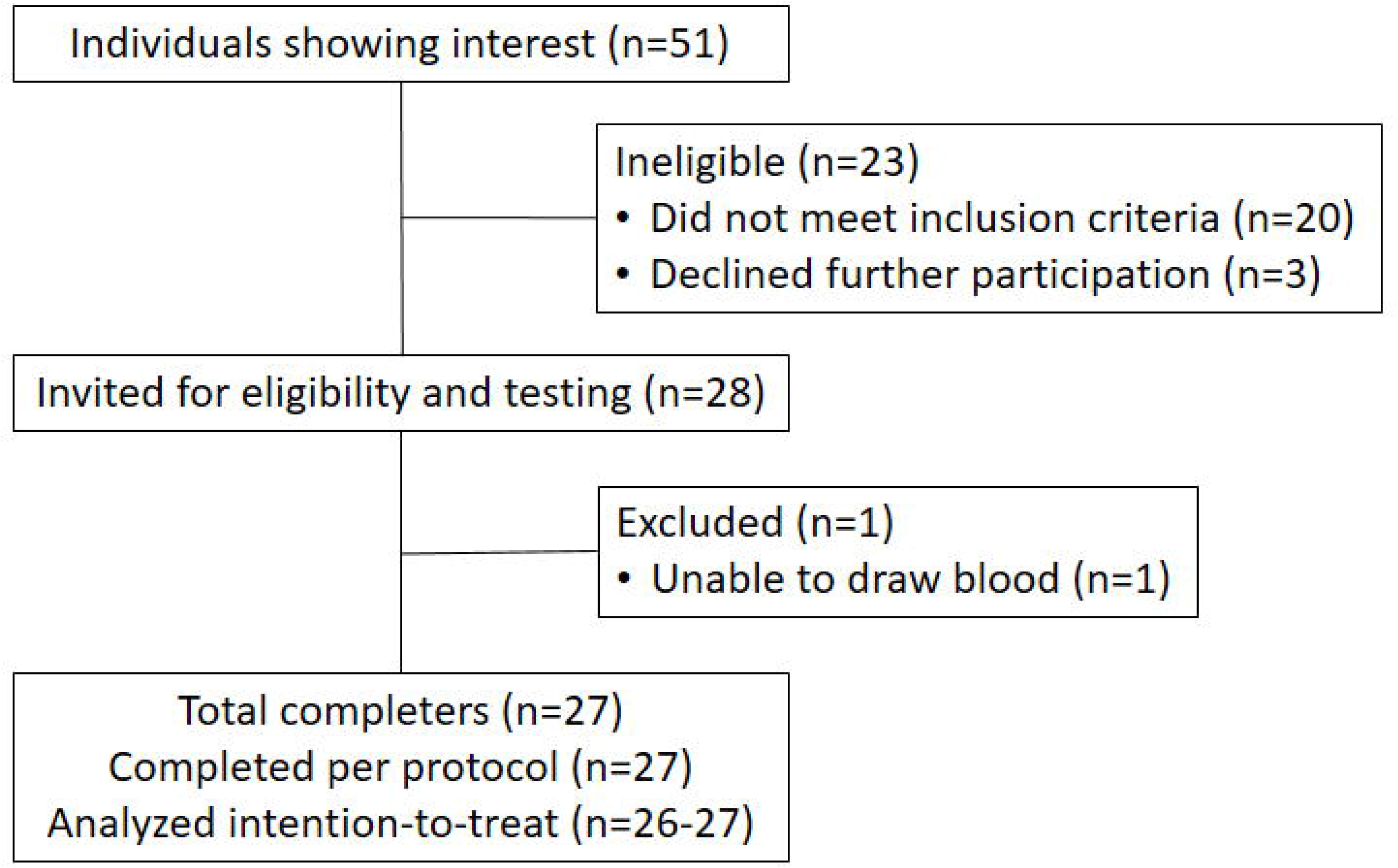
CONSORT flow diagram. CONSORT, Consolidated Standards of Reporting Trials.

Subjects consumed 404.9±55.3 mg of caffeine (CI 95% 382 – 427.7) before the test, rising to plasma 0.086±0.04 nmol/L (CI 95% 0.06 – 0.11) after 1 hour. Even after 48 h of withdrawal of 23 caffeinated foods and drinks, plasma levels were 0.011±0.03 nmol/L (CI 95% 0 – 0.04) when treated with a vehicle. We followed the subjects for 24-hours and reported no adverse effects such as tachycardia, angiogenesis, insomnia, gastrointestinal discomfort, or other symptoms.

### Caffeine increases fatigue resistance of spinal muscles

*Primary outcome*. Caffeine had a small effect size on increasing 13.8±6.3 % Biering-Sørensen endurance, but not peak force (Table 2).

Secondary outcomes. Caffeine induced a moderate effect size on increased blood lactate production (Table 2), a biomarker of exercise intensity. Biering-Sørensen test reached RPE intensities classified as moderate to intense (F_6,258_ = 64, *η*^2^ = 0.6, β = 1.0, P<0.05,Fig. 2A), Moreover, caffeine had a moderate effect on decreased RPE (Table 2). HR increased during the test (F_13,637_ = 6, *η*^2^=0.11, β=1.0, P<0.05, Fig. 2B), and caffeine demonstrated a great positive chronotropic effect during the physical test (P< 0.05, Fig. 2C, Table 2).

**Fig. 2.**
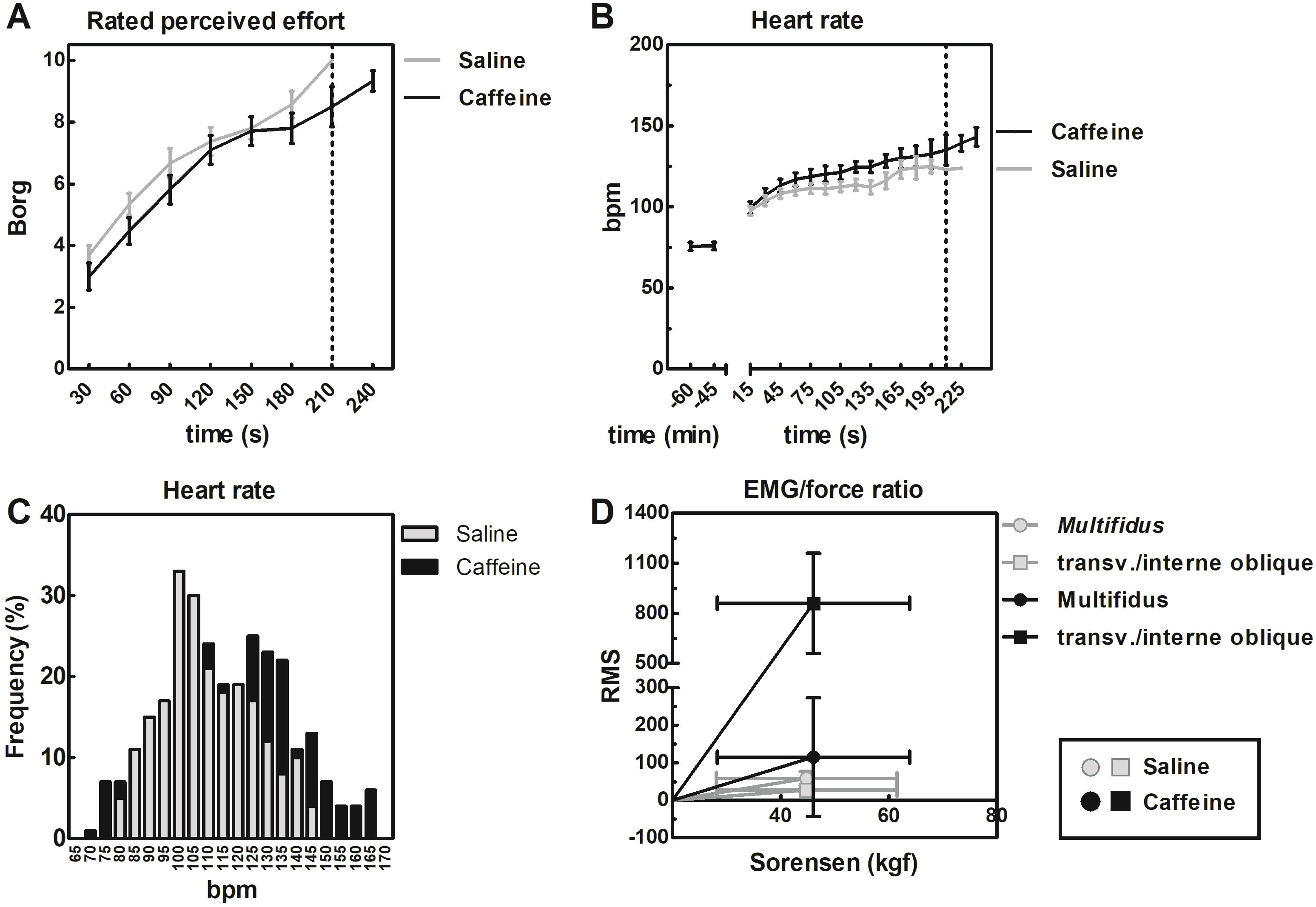
Effects of caffeine on rated perceived effort (A), heart rate (B-C) and EMG/force ratio during the Biering-Sørensen test.

### Caffeine stimulates spinal muscles activity

#### Secondary outcomes

The EMG-force ratio determines the neuromuscular status of muscle. The EMG-force ratio shows that caffeine decreases fatigue (right shift) in the multifidus spinae and transversalis/interne oblique muscles (Fig.2D).

EMG revealed that caffeine increased muscle recruitment and frequency of stimulation of multifidus spinae and abdominal, suggesting attenuation of muscle fatigue. The median frequency of multifidus spinae decreased over Biering-Sørensen test (F_29,1131_ = 127, *·^2^* = 0.76, P<0.05, Fig.3A), an EMG marker of muscle fatigue, without differences in treatment (F_1,39_ = 0.2, *η^2^* =0.005, P<0.05, Fig.3A). However, the distribution of median frequencies revealed higher frequencies on caffeine-treated subjects (χ^2^_1_=144, P<0.05, Fig.3B). Caffeine still had a large effect size (*η^2^*=0.49) on increasing multifidus spinae RMS values (recruitment of new muscle fibers) throughout the Biering-Sørensen test, from the beginning (F_1,540_ = 53, P<0.05,Fig. 3D) to the end of the test (P< 0,05, Fig. 3E).

**Fig. 3.**
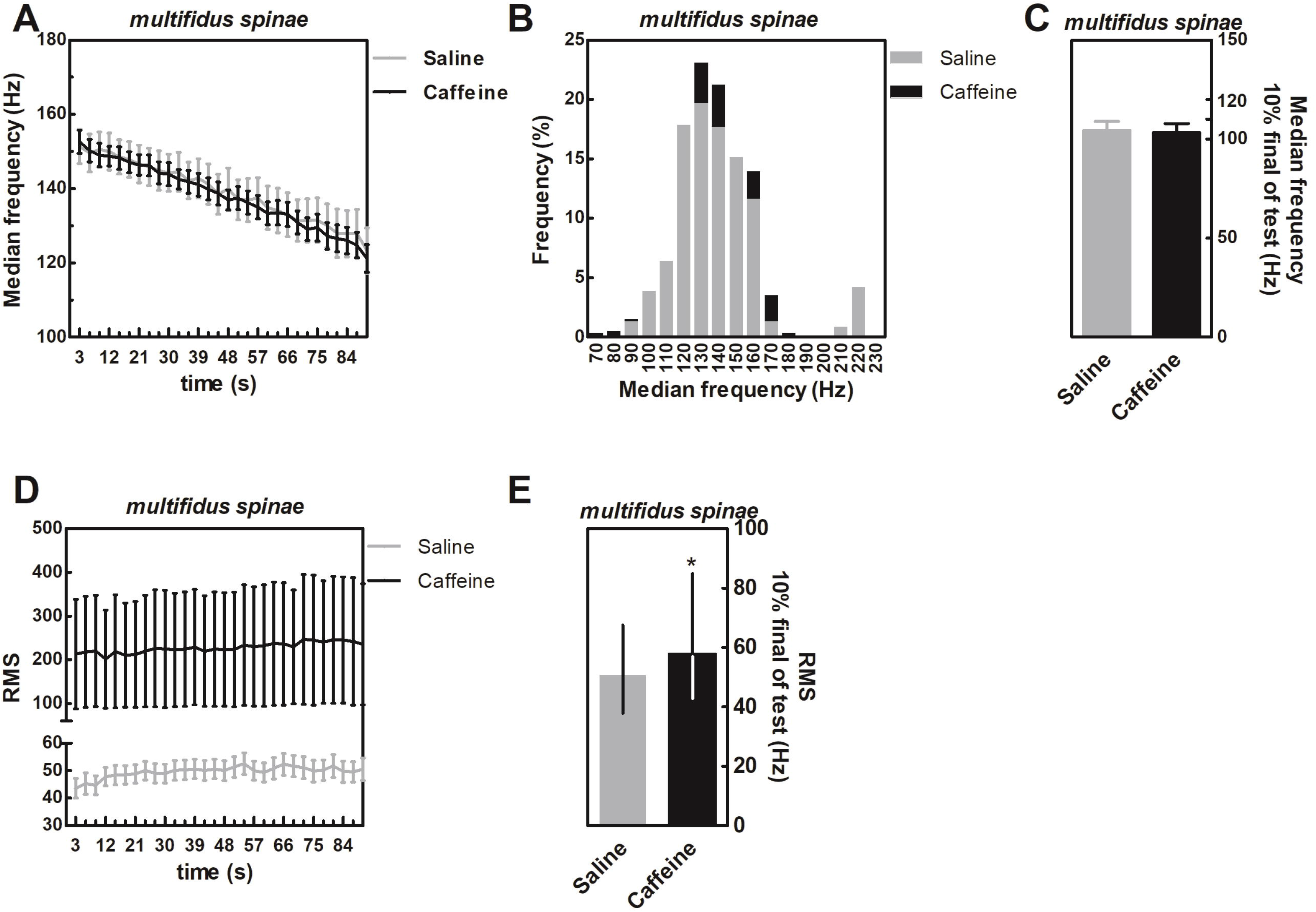
Effects of caffeine on the median frequency (A-C) and recruitment (D-E) of the *multifidus* spinae muscle during the first 90 seconds and 10% final of the Biering-Sørensen test. * P< 0.05 vs. saline (Wilcoxon matched-pairs signed-rank test).

Caffeine ergogenic effect size was medium (*η^2^* = 0.13) on increased median frequency (F_29,1073_ = 9.7, P<0.05, Fig. 4A) of transversalis/interne oblique. The distribution of median frequencies shifted to the right, for higher frequencies (*η*^2^_2_ = 2900, P<0.05,Fig. 4B) as in the multifidus spinae. The effect size of caffeine-increased muscle recruitment was also large (*χ^2^* = 0.94, F_1,540_=87, P<0.05,Fig. 4D). These ergogenic effects disappeared in the final 10% of the Biering-Sørensen test (Fig.4C, and E).

**Fig. 4.**
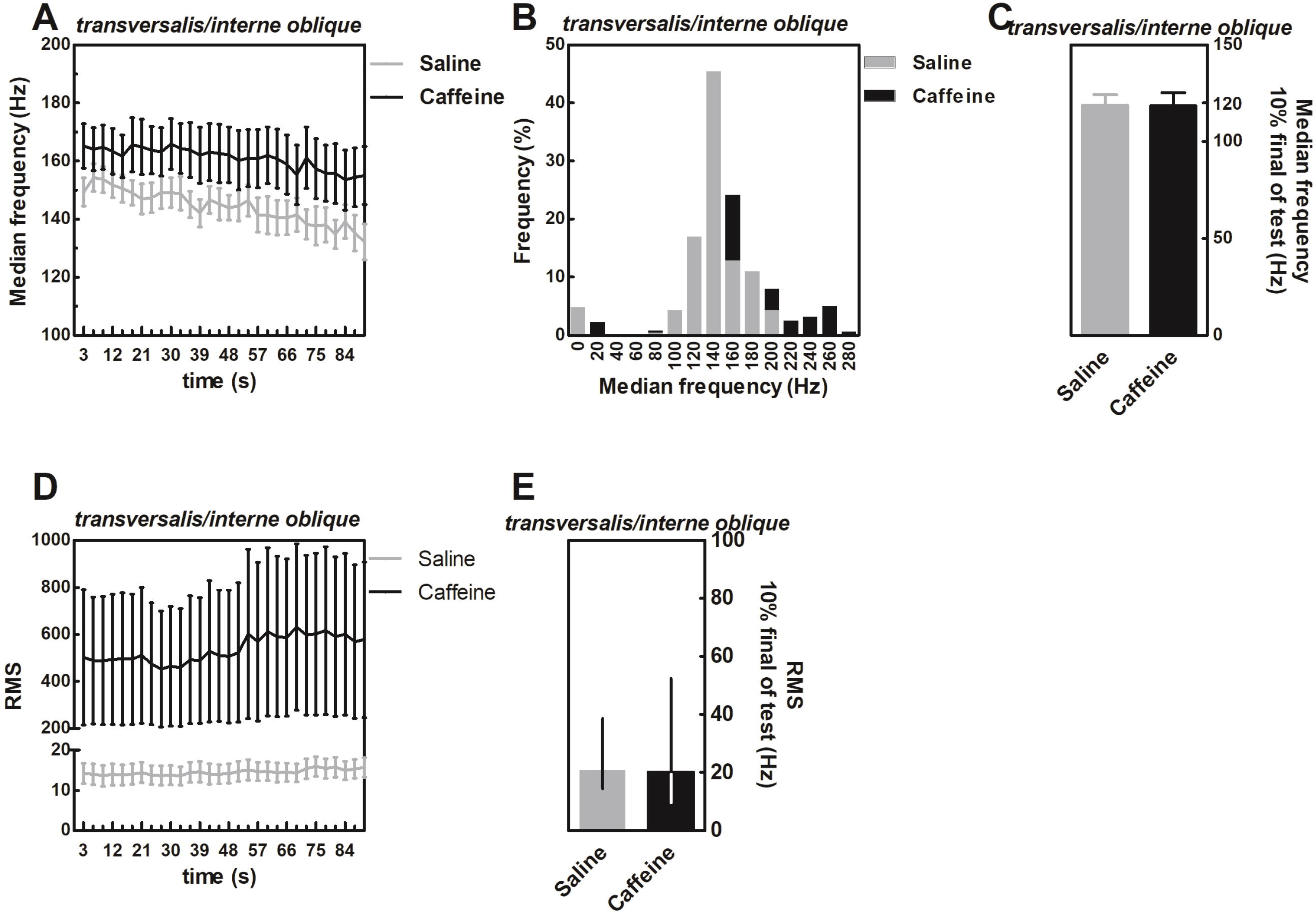
Effects of caffeine on the median frequency (A-C) and recruitment (D-E) of the transversalis/interne oblique muscles during the first 90 seconds and 10% final of the BieringSørensen test.

## DISCUSSION

### New ergogenic effects of caffeine on postural muscles

We first demonstrated the ergogenic effects of caffeine on postural muscles – *multifidus spinae* and *transversalis*/interne oblique, components of core muscles. Core muscles are essential for physical performance and injury prevention, goals of sports medicine. The increase Biering-Sørensen endurance (or resistance to fatigue) had small effect size, as well established for other muscle groups and activities [4–6,9]. The Biering-Sørensen test is the most robust predictor for low back pain and trunk stability [43], with implications from health to elite sport. Strengthening core muscles increases Biering-Sørensen test performance and decreases low back pain [43,44]. There is not much evidence in the sports sciences. Water polo, soccer, and rowing athletes performed well in the Biering-Sørensen test (163.6±50.7 sec) [41,42]. Muscle activity of abdominal and spine extensors of long-distance runners and triathletes were higher in the Biering-Sørensen test, which contributes to running performance, such as more excellent absorption by the trunk muscles of disrupting torques generated by the lower limbs [41,42].

The Biering-Sørensen test performance also directly influences athletes’ motivation and self-efficacy [56]. Athletes are more resistant to high blood lactate concentrations, pain, and fatigue [2,7]. Psychological factors are associated with this performance, such as improved motivation and experience [4–6,9,57]. Moderate doses of caffeine (3–6 mg/kg) decrease RPE in athletes [4–6,9]. We observed this same effect of caffeine (6 mg/kg) in this spine muscle stress test. Effort perception is a brain function; neurophysiology involves central fatigue and diminished cortical arousal [58], increased cortical activity during movement in the primary motor and supplementary movement areas [59], and increased activation of the temporal and insular cortex [60]. This evidence retrieves the CNS’s role in the development of central fatigue [24,27,28,33–35,57,61–64]. Here, the effect of caffeine on RPE and EMG reinforces this idea.

### Caffeine decreases central fatigue

Caffeine modified the EMG response in the Biering-Sørensen test. EMG activation is the preliminary condition for any force development. Here, caffeine-induced a substantial increase in muscle recruitment, a moderate increase in the high frequency of stimulation, and decreased fatigue (EMG/force ratio) of *multifidus spinae* and *transversalis*/interne oblique muscles. There is no evidence about the effects of caffeine on these muscles, but other muscle groups have a similar EMG response to caffeine. Caffeine (5 mg/kg) improved performance in the cycling time trial and increased Maximal voluntary strength (MVC), power output, and muscle recruitment of vastus *lateralis and medialis* [38,40]. Caffeine (6 mg/kg) increased the elbow’s maximum flexor torque, muscle recruitment, and fiber conduction velocity [40]. Muscle strength was higher in electrically stimulated *adductor pollicis* and ankle dorsiflexors muscles of caffeine-supplemented subjects (500 mg and 6 mg/kg) [37,39]. Caffeine decreased low-frequency fatigue (20, 30, and 40 Hz stimulation) in these muscles, which is a reduction in expected muscle force (fatigue) due to an impairment of excitation-contraction coupling. Caffeine did not modify muscle strength at higher stimulation frequencies (> 40 Hz) [37,39], such as those achieved voluntarily in this study.

This evidence reinforces the role of caffeine in central fatigue, which also modifies cortical silent period (CSP) and spinal excitability [27,28]. A transcranial magnetic stimulation (TMS) fatigued abductor digiti minimi muscle and caffeine reduced CSP [27]. CSP refers to an interruption of voluntary contraction by electrical or magnetic stimulation of the motor cortex. Caffeine also increased spinal excitability by increasing the slope of the H-reflex recruitment curve normalized to M wave (H_slp_/M_slp_), which provides an alternative measure of monosynaptic reflex gain [28]. These are evidence of the neurophysiological components of the ergogenic effects of caffeine. Moreover, our experimental design removed caffeine for 48 hours from subjects, moderate coffee consumers, which may have enhanced our ergogenic results [65].

### Advancing caffeine’s mechanisms of action: negative allosteric neuromodulation of adenosinergic receptors

However, the mechanisms of action of caffeine’s ergogenic effects are intriguing. For a long time, literature insisted on the mobilization of intracellular Ca^+2^ and inhibition of phosphodiesterase [1,2,7,66]. These biological effects are achieved *in vitro* (in millimolar concentrations, 10^−3^), but toxic and lethal *in vivo* [12,67]. Recent studies discuss CNS’s antagonism of the adenosinergic system as a possible candidate mechanism of caffeine [27,28,33,34]. Electrophysiological studies support the positive effect of caffeine on vigilance, attention, speed of reaction, information processing, and arousal [27]. Caffeine acts on the CNS decrease the perception of effort and modifies the motor drive. A1 receptor (A_1_R) and A_2A_R are primarily responsible for the central effects of adenosine [68], also the main target of nontoxic psychostimulant doses of caffeine in micromolar levels (10-625) [67,69]. Generally, blocking presynaptic A_1_R increases the probability of neurotransmitter release, including dopamine and serotonin, whereas blocking presynaptic A_2A_R decreases neurotransmitter release [17,68,70]. A1 blockade could reinforce Meeusen’s untested hypothesis about central fatigue [62], an exercise-induced increase in extracellular serotonin stimulating sleep, lethargy, and drowsiness, and loss of motivation. Adenosine is a neuromodulator responsible for these latter effects [71,72]. Caffeine decreases CSP in fatigued muscles [27]. Experimental manipulation of GABA_B_ inhibitory presynaptic receptors modulate CSP [73,74]. In this context, caffeine might interfere with GABAergic neurotransmission in different ways [75]. Moreover, basal forebrain cholinergic neurons are under tonic inhibitory control of endogenous adenosine [71]. A_2A_R is highly expressed in the basal ganglia, with emphasis on the ventral striatopallidal GABA pathway, where they form functional heteromeric receptor complexes (A_2A_R-D_2_) with inhibitory presynaptic dopaminergic D_2_-like receptors. D2-like receptors decrease adenylyl cyclase activity through G_i/o_ proteins [76]. Caffeine and selective A_2A_R antagonists produce psychostimulant effects, not just by competing with adenosine for its binding to the A_2A_R, but also by exerting a negative allosteric modulation within the A_2A_R of A_2A_R-D_2_ heterotetramer [76]. Under normal conditions, employing the negative heteromeric allosteric modulation, endogenous adenosine tonically inhibits psychomotor activation mediated by tonic activation of D_2_R by endogenous dopamine. Caffeine, through negative allosteric modulation, counteracts the effect of adenosine and produces psychomotor activation [76]. Caffeine (a non-selective A_1_ and A_2A_R antagonist) and SCH 58261 (potent and selective A_2A_R antagonist) are psychostimulants [33,77,78]. Davis [34] demonstrated that NECA (A_1_R and A_2A_R agonist) inhibits the ergogenic effects of caffeine in rats. We demonstrated that SCH 58261 is ergogenic in mice and that the ergogenic effect of caffeine disappears in mice knocked out to neuronal A_2A_R in the CNS [33]. In the present study, we demonstrated that caffeine decreases the perception of effort and increases muscle excitation in humans, increasing the excitation-contraction efficiency of spine muscles. This evidence jointly reinforces the role of caffeine in mitigating central fatigue during exercise.

### Health impacts

Caffeine supplementation did not impact the subjects’ general health. Monitoring the subjects for 24 hours did not reveal any known adverse effects of caffeine, such as tachycardia, angiogenesis, insomnia, gastrointestinal discomfort, or other symptoms. Dietary consumption and experimental treatment offered approximately 400 mg of caffeine daily to the subjects, a dose considered safe and free of adverse effects in moderate consumers [18].

### Limitations

We have identified two limitations of this study. We did not perform a blinding assessment and identified residual caffeine in the plasma of some subjects. This last point shows flaws in the withdrawal protocol. However, blood caffeine concentrations are not a standard measure in most studies focusing on ergogenicity. At the same time, blood caffeine is a strong point of the study. This study is a simple and reproducible experimental design. Increasing the sample size to include patients and athletes of different sexes and ages is necessary for generalization and applicability of these results.

## CONCLUSION

In summary, our results show that caffeine is ergogenic for postural muscles. As an ergogenic resource, caffeine may benefit patients with back pain. The results also reinforce the ergogenic role in sport. Decreased RPE and improved muscle activity suggest central mechanisms of caffeine. That is, caffeine attenuates central fatigue during acute exercise.

## Data Availability

Anonymous data will be available beginning three months and ending five years following article publication for researchers who provide a methodologically sound proposal.

## Acknowledgements

We thank Dr. Fernando Diefenthaeler (Biomechanics Laboratory/CDS/UFSC) for his assistance with EMG routines and Dr. Alexandra Latini (LABOX/CCB/UFSC) for using HPLC.

## Contributors

LC, IJCS, HK, and ASAJr conceived the study and designed the study in collaboration. LC, TN, MG, MCS, and VB carry out recruitment, blinding, experiments, and data collection. ACBA and DLS performed the HPLC. LC and ASAJr analyzed the data. ASAJr did the statistical analysis. LC, TR, and ASAJr wrote the manuscript. All authors interpreted the data and contributed to subsequent drafts of the manuscript, and all authors have seen and approved the final version.

## Ethics approval

was obtained (CAAE 86400418.0.0000.0121) from the Brazilian human ethics committee (http://plataformabrasil.saude.gov.br).

## Trial

The Universal Trial Number (UTN) is U1111-1234-2079. The protocol consultation is available in the Brazilian Registry of Clinical Trials (http://www.ensaiosclinicos.gov.br/) under number “RBR-39JNP4” and title “The biology of the ergogenic effects of caffeine.”

## Data sharing statement

Anonymous data will be available beginning three months and ending five years following article publication for researchers who provide a methodologically sound 23 proposal.

## Provenance and peer review

Not commissioned; externally peer-reviewed.

